# Vitamin D for painful diabetic neuropathy: a systematic review and meta-analysis of randomised controlled trials

**DOI:** 10.1101/2025.03.10.25323653

**Authors:** Abraham Gilbody, Joseph Gilbody

## Abstract

**Background:** Diabetes mellitus contributes increasingly to the Global Burden of Disease [GBD]; particularly in high and increasingly in low-and middle-income countries. Strategies to prevent and mitigate the impact are a public health priority. Painful diabetic neuropathy (PDN) is a syndrome of sensory disorders caused by both type 1 and type 2 diabetes mellitus. Available treatments include antidepressant medications and strong analgesics. These are often only partly effective and associated with significant side effect profiles. There is a need for effective treatments with low toxicity. Vitamin D has been proposed as potential therapeutic and biologically plausible agent.

Non-randomised studies demonstrate benefit, but are subject to biases. There is a need for robust evidence derived from randomised data to inform patient care in this debilitating complication of diabetes.

**Review aim:** To synthesise randomised controlled trials (RCTs) of Vitamin D supplementation and its effects on painful diabetic neuropathy.

**Review methods:** A range of databases [Medline, EMBASE, Web of Science, Cochrane Library, CINHAL, EBSCO and Google Scholar] were searched from inception to March 2025, with backwards and forward citation searches to identify eligible studies. RCTs comparing Vitamin D with placebo in patients with diabetes [type 1 or 2] and PDN were sought. The primary outcome was pain as measured using a validated pain measure or measure of PDN. A fixed effects meta-analysis of continuous pain data was conducted, with standardisation between studies to calculate a standardised mean difference [SMD] between Vitamin D and placebo. Small study and publication bias was tested using an Egger plot, and study quality was assessed using the Cochrane Risk of Bias [RoB] tool.

**Results:** Four eligible studies were identified and three of these studies [comprising 260 participants] provided meta-analysable data. There was a statistically significant short-term benefit for vitamin D (pooled standardised mean difference =-0.70; 95%CI-0.95 to-0.45). There was moderate between study heterogeneity, and there was an intermediate level of heterogeneity (I = 54.9%). No studies reported medium-or long-term outcomes. The quality of studies was variable (either low or moderate risk of bias), with poor concealment of allocation as the most important design limitation.

Two of the studies had been prospectively registered, making it difficult to check for bias in one study due to potential selective reporting of outcomes. Despite conducting an Egger Funnel Plot, it was not possible to exclude the influence of small study and selective publication bias.

**Discussion:** Based on a meta-analysis of non-registered small size studies there was evidence of short-term reduction in pain symptoms in painful diabetic neuropathy. This benefit needs to be confirmed in fully powered RCTs with a longer duration of follow up. Vitamin D remains a viable low-cost treatment option for PDN, but more research is needed before this can confidently be recommended for routine patient care.

## INTRODUCTION

Painful diabetic neuropathy (PDN) is a syndrome of sensory disorders caused by both type 1 and type 2 diabetes mellitus. PDN is known to be not only one of the most common diabetic peripheral neuropathies, but is also detrimental to the overall health of people with diabetes [1].

PDN is characterised by distressing tingling, burning, sharp, shooting, and lancinating or even as electric shock sensations [2], with nocturnal exacerbation and impacts on sleep. PDN also contributes to diabetic foot disease by causing poorly-healing ulcers and eventual amputation [3]. This contributes to reduced physical activity, depression, and decline in quality of life (QOL) [4].

Epidemiological data demonstrates that the incidence of pain symptoms in diabetes is 10-20% and 40-50% with diabetic neuropathy [5]. PDN also contributes to higher rates of healthcare utilisation and costs [6]. Patients with PDN have increased costs due to more frequent and prolonged hospitalizations and increased rates of outpatients visits, coupled with reduced economic productivity and sickness absence [6].

Treatment of PDN comprises three approaches: (1) intensive glycaemic control and risk factor management, (2) treatments based on pathogenetic mechanisms, and (3) symptomatic pain management [7]. Intense glucose control is currently the only intervention that has been proven to reduce the risk of development of neuropathy [8, 9]. Peripheral nerve injury is largely understood as the main source of pain, however recent literature supports the role the central nervous system in the disinhibition and amplification of pain [10].

Consequently, clinical guidelines recommend symptomatic relief of neuropathic pain through the ‘centrally acting’ analgesic effects of antidepressant drugs (tricyclic antidepressants or selective serotonin reuptake inhibitors), anticonvulsants, and opioids [11]. Treatment of PDN is far from satisfactory with many medications providing limited pain relief with a significant side effect profile [7].

Another treatment modality is emerging is the use of Vitamin D which will be the focus of this systematic review, and meta-analysis. Vitamin D deficiency been shown to be an independent risk factor for the development of PDN [12]. Vitamin D has also been shown in some studies to have a significant influence in PDN compared to painless DPN [13]. Furthermore, non-randomised evidence shows that high-dose intramuscular injections of vitamin D can reduce pain [14]. This all presents the biologically-plausible mechanism that the correction of low vitamin D might in turn have therapeutic benefit in PDN. However definitive evidence will only emerge from robust controlled clinical studies.

Research intelligence is needed to establish whether vitamin D therapy treats or delays the onset of symptoms in painful diabetic neuropathy.

## REVIEW AIM

The overarching aim of this systematic review was to synthesise data from randomised controlled trials of Vitamin D supplementation and its effects on painful diabetic neuropathy.

By using systematic review methods, a supplementary aim was to map the strengths and limitations of existing available research, and to make recommendations for clinical practice and identify the need for further research.

## REVIEW METHODS

The systematic review was undertaken in march 2025 in accordance with the Methods of the Cochrane Handbook for Systematic Reviews [15], and followed the Preferred Reporting Items for Systematic Reviews and Meta-Analyses (PRiSMA) guidelines [16].

### Search strategy

A search strategy was devised to identify relevant studies from the following electronic databases: Medline, EMBASE, Web of Science, Cochrane Library, CINHAL, EBSCO and Google Scholar. These databases were selected, as recommended by the Cochrane Handbook, as MEDLINE, Cochrane Central and Embase are effective in identifying randomized controlled trials [15]. The additional databases facilitates a wider and more extensive search of sources thus minimizing selection bias for the studies [15]. Search terms were devised from the PICO structure [described below] such as ((diabetic AND neuropathy AND painful AND (“25(OH) D” OR “25-Hydroxyvitamin D” OR “vitamin d”)). The search utilized optimal Cochrane strategies to identify randomized controlled studies [17], whilst excluding non-randomised data (a sensitive and specific search). Resources were only available for English language publication, therefore the search was restricted.

In addition to the electronic bibliographic searches, reference lists from included papers were scrutinised to identify additional studies [‘backwards citation searching’]. Additional studies citing included studies were also sought using Google Scholar [‘forward citation searching’] [18].

### Criteria for study selection

Only randomised controlled trials (RCTs) were considered as they are considered the gold standard design for studying cause-effect relationships between an intervention and outcome. Randomization is the optimum design in eliminating much of the bias inherent with other study designs, and [most importantly] minimises the distortion of cause-effect from both known and unknown confounding factors [19]. Non-randomised controlled studies, reviews and case-series were excluded. Potentially eligible studies were assessed for inclusion against the PICO criteria for inclusion in the systematic review are summarised in table 1.

**Table 1:**
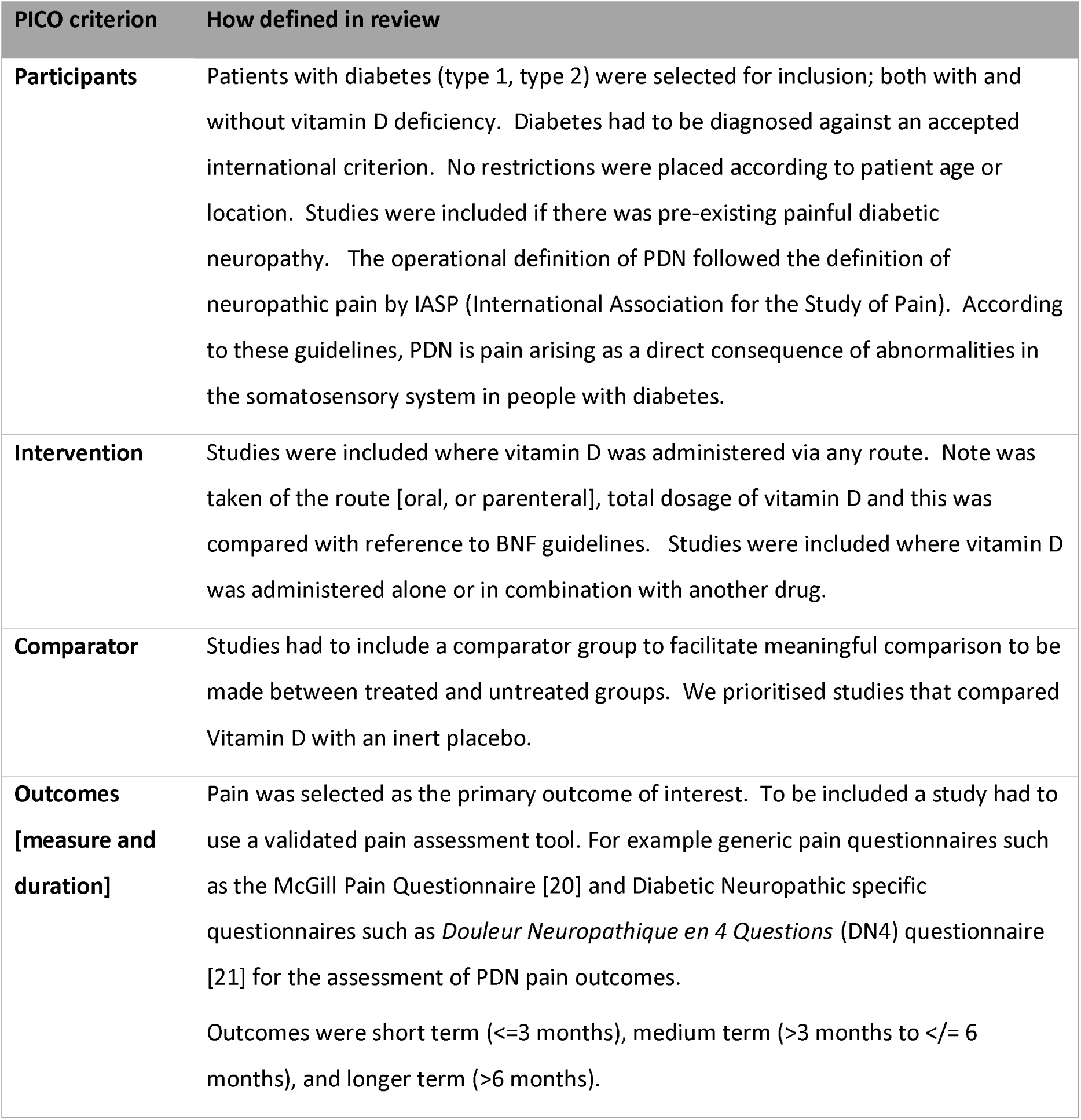
study inclusion criteria for systematic review.

| PICO criterion | How defined in review |
| --- | --- |
| **Participants** | Patients with diabetes (type 1, type 2) were selected for inclusion; both with and without vitamin D deficiency. Diabetes had to be diagnosed against an accepted international criterion. No restrictions were placed according to patient age or location. Studies were included if there was pre-existing painful diabetic neuropathy. The operational definition of PDN followed the definition of neuropathic pain by IASP (International Association for the Study of Pain). According to these guidelines, PDN is pain arising as a direct consequence of abnormalities in the somatosensory system in people with diabetes. |
| **Intervention** | Studies were included where vitamin D was administered via any route. Note was taken of the route [oral, or parenteral], total dosage of vitamin D and this was compared with reference to BNF guidelines. Studies were included where vitamin D was administered alone or in combination with another drug. |
| **Comparator** | Studies had to include a comparator group to facilitate meaningful comparison to be made between treated and untreated groups. We prioritised studies that compared Vitamin D with an inert placebo. |
| **Outcomes**<br>**[measure and**<br>**duration]** | Pain was selected as the primary outcome of interest. To be included a study had to use a validated pain assessment tool. For example generic pain questionnaires such as the McGill Pain Questionnaire [20] and Diabetic Neuropathic specific questionnaires such as *Douleur Neuropathique en 4 Questions* (DN4) questionnaire [21] for the assessment of PDN pain outcomes.<br>Outcomes were short term ($\leq 3$ months), medium term ($> 3$ months to $\leq 6$ months), and longer term ($> 6$ months). |

### Data collection and extraction

Relevant data were first extracted into a spreadsheet. Relevant data were: the mean pain scores, % change in pain score, Dosage and type of Vitamin D supplementation used, standard deviations, sample sizes and patient gender in both intervention and control groups. Study authors were contacted to provide missing data.

### Quality assessment of trials

As per the Cochrane Handbook (21), trials were assessed on quality based on 3 main parameters (greater detail in appendix 1):

- Randomisation (Allocation sequence generation and allocation sequence concealment present)
- Blinding (Double blinding, Effective blinding, Bias due to measurement of outcomes, Bias due to missing outcome data)
- Attrition bias (Loss to follow up, correct analysis preformed)

Each parameter underwent a risk-of-bias judgement and was assigned to 3 groups:

‒ **Low** risk-of-bias
‒ **Moderate** risk-of-bias
‒ **High** risk-of-bias

The overall judgement of bias was based on the collection of ratings. Quality assessed data were plotted into the risk of bias (RoB) tool found on Revman5 [see table 2]. This presented the overall judgement of risk of bias using an infographic. The GRADE tool was used to assess the quality of the data from each study using several domains. Study quality was marked down for: Risk of bias, Imprecision, Inconsistency, Indirectness and Publication bias. Marked up for: Magnitude of effect, Dose-dependent gradient and all residual confounding would decrease magnitude of effect The criteria for Cochrane for randomisation [Cochrane Handbook for Systematic Reviews of Interventions 4.2.5(21)] are reproduced in appendix 1.

**Table 2:** Overall risk-of-bias judgement criteria.

| **Overall risk of bias judgement** | **Criteria** |
| --- | --- |
| Low risk of bias | The study is judged to be at **low risk of bias for all domains** for this result. |
| Moderate risk of bias | The study is judged to raise **moderate risk of bias** in at least one domain for this result, but not to be at **high risk of bias** for any domain. |
| High risk of bias | The study is judged to be at **high risk of bias** in at least one domain for this result.<br>Or<br>The study is judged to have **moderate risk of bias** for multiple domains in a way that substantially lowers confidence in the result. |

### Small study, publication and outcome reporting bias

A range of databases were searched to minimise study identification and selection biases. Publication bias arises through the inclusion of only published studies, and the potential omission of negative studies since these are often unpublished [15]. A funnel plot analysis was planned to detect if authors publish smaller studies with positive results [15]. This is commonly found among studies which are smaller in size which are typically performed with reduced methodological rigor producing an overestimation of the intervention effect [15]. An asymmetrical funnel plot indicates that smaller studies were subject to publication and small study bias. The greater the asymmetry the increased likelihood that the amount of bias will be substantial. To quantify the magnitude of funnel plot asymmetry, an Egger test was planned [22].

Selective reporting of outcome bias was addressed (where possible) by comparing the outcomes described in the study trial register or pre-study protocol with the range of outcomes reported in the final study publication [23].

### Proposed meta-analysis

In line with Cochrane guidance, where similar [non-heterogenous] data were available for three or more studies, then a fixed effects meta-analysis was undertaken using RevMan 5 software [24]. It was anticipated that the primary pain outcome would be presented as a continuous variable of change in pain scores in both control and intervention groups. Endpoint data in each group were combined to calculate between-group differences. Where trials used different instruments to measure the pain outcomes a standardized mean difference (SMD) with 95% confidence intervals was calculated. The standardised mean difference measures the size of the intervention effect of each study compared to the between-participant variability in outcome measurements recorded in each individual study. To allow for easier interpretation and analysis of the results, we planned to convert the SMD into the specific clinical measures used in the studies (such as the McGill Pain Questionnaire, or the Leeds Assessment of Neuropathic Symptoms and Signs (LANSS) scale). This can be done by multiplying the SMD by an estimate of the standard deviation (SD) associated with the outcome measure instrument, according to the Cochrane Handbook.

### Assessment of heterogeneity

Heterogeneity was tested using Higgins’ test (I) to test for between-study variation in order to determine if studies can be pooled for meta-analysis [25].

- If <75% use fixed effects
- If more than 75% use random effects
- If more than 90% just use systematic narrative review (no meta-analysis undertaken)

Where significant heterogeneity was found, sources of heterogeneity were explored. Anticipated source of heterogeneity included: dose and route of Vitamin D; baseline Vitamin D status of participants, duration of follow up.

## RESULTS

### Study selection

The selection process of summarised using the PRISMA flow diagram (figure 1). The literature search identified a total of 73 potentially eligible studies. After removal of duplicates, 63 papers were included in the initial screening procedure. Electronic titles and abstracts of papers from the search were first screened for eligibility. Following the removal of 12 papers, the remaining papers were screened on their full text against PICO criteria detailed below. Where the full text was not available study corresponding author was contacted via email. In 4 papers were found the be eligible for use in the systematic review [26–29] and three provided sufficient data for the meta-analysis [26] [27, 28]. Common reasons for exclusion included: inappropriate outcome measurements, lack of comparator groups and inappropriate study design (non-randomised).

**Figure 1:**
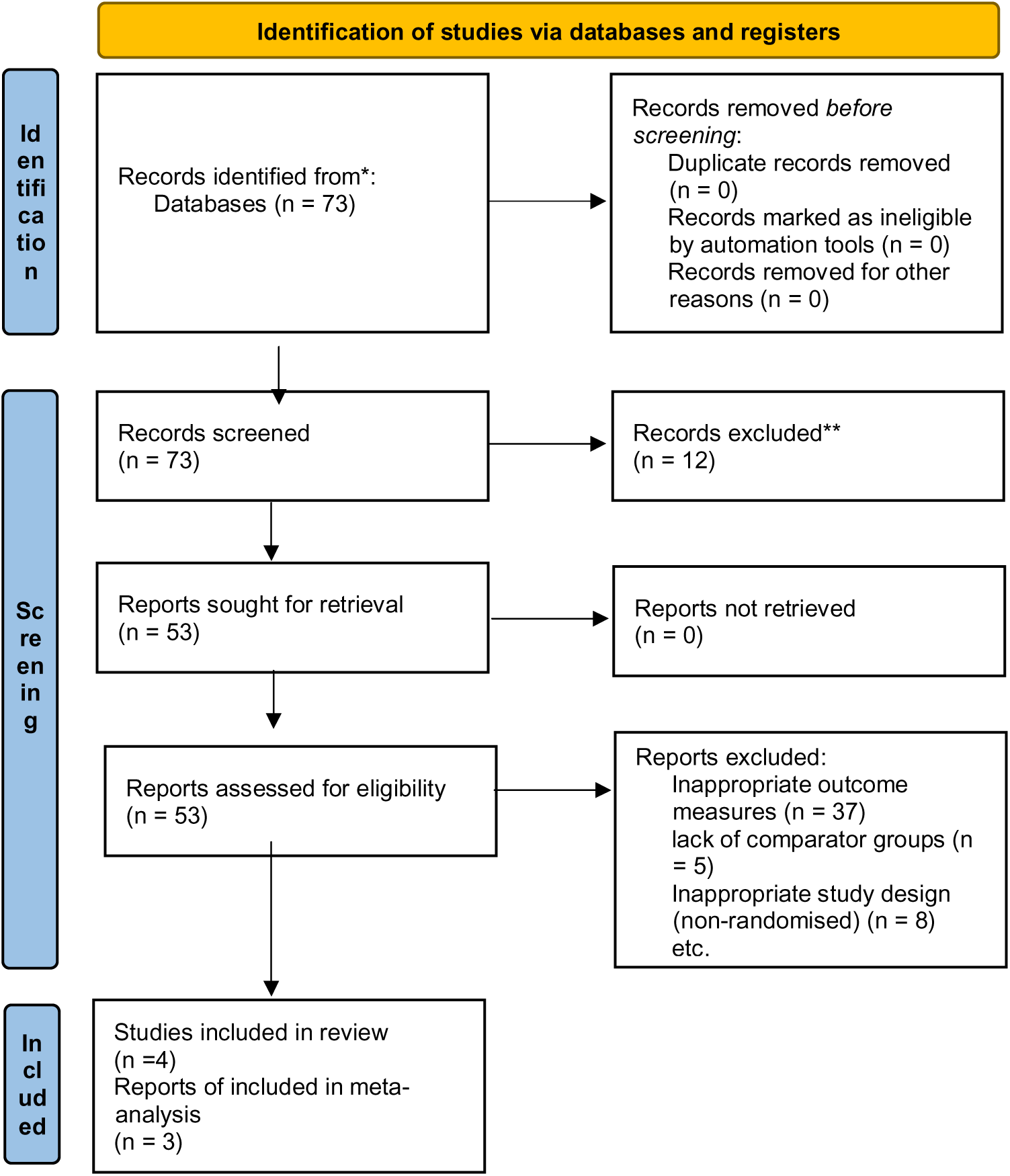
PRISMA flow diagram

Three studies that were randomised and met the PICO inclusion criteria [26] [27, 28] were eligible for inclusion into the meta-analysis. One study was a five-arm trial, and two arms provided data that allowed a comparison between Vitamin D and placebo. Study size ranged from 68 to 112 and 260 participants were eligible for meta-analytic pooling. The weighting of each paper in the meta-analysis was proportional to their respective participant totals (and inversely related to the variance). Each study reported a different pain scales, with the visual analogue scale [28], the NSS [26] and using the Neuropathic pain scale (NPS) [27]. Each study recruited patients who were Vitamin D deficient at baseline, with placebo arm levels ranging from 11.68 ng/mL (SD 3.8) to 24.6 ng/mL (SD 5.5) [27]. Vitamin D was administered orally in all cases, and dose regimens ranged from 4,000 IU per day to 50,000 IU over two weeks. Duration of follow up ranged from 8 weeks [28] [26] to 12 weeks [27]. For PICO and study details see table 3.

**TABLE 3:** DATA EXTRACTED FOR STUDIES MEETING PICO INCLUDING CRITERIA.

| STUDY | Design | Participant characteristics | Pain outcome | Intervention | Comparator |
| --- | --- | --- | --- | --- | --- |
| **Shehab et al (2015)** | Placebo-controlled randomised trial | N = 112 participants (Placebo = 55, Vitamin D = 57)<br>Sex: Placebo (M 27 F 28)<br>Vitamin D (M 21 F 36)<br>Age: Placebo (59.8 years SD = 9.6)<br>Vitamin D (61.8 SD = 8.1)<br>Baseline Vitamin D: Placebo (29.2 nmol/mL SD 9.5) (11.68 ng/mL SD 3.8)<br>Vitamin D (25.3 nmol/mL SD 10.9) (10.12 ng/mL SD 4.36)<br>Final Vitamin D level: Placebo (30.3 nmol/mL SD 8.9) (12.12 ng/mL SD 3.56)<br>Vitamin D (58.2 nmol/mL SD 23.8) (23.28 ng/mL SD 9.52)<br>Change in Vitamin D level: Placebo (1.1 nmol/mL SD 3.6) (0.44 ng/mL SD 1.44)<br>Vitamin D (32.8 nmol/mL SD 23.7) (13.12 ng/mL SD 9.48)<br>Period of follow up: 8 weeks | Primary outcome<br>Neuropathic Symptom Score (NSS)<br>Placebo: pre-test (5.50 SD = 1.25) after test (5.45 SD = 1.20) Difference (-0.20 SD 0.59)<br>Treatment: pre-test (5.92 SD 1.29) post-test (4.43 SD 1.58) Difference (-1.49 SD 1.37) P<0.001<br>Outcome measured at 8 weeks | The treatment group received oral capsules of vitamin D3 (a capsule of cholecalciferol, 50,000 IU, Bronson, Lindon, Utah, USA) once weekly for 8 weeks. | The placebo group received starch capsules, also once weekly for 8 weeks. |
|  |  | Diagnosis: Adult patients with type 2 diabetes with DPN and vitamin D deficiency |  |  |  |
| **Pinzon et al (2021)** | Randomized controlled clinical trial | N = 68 (placebo = 34 Vitamin D = 34)<br>Sex: Female 41 (60.3%)<br>Male 27 (39.7%)<br>Age: mean: 64.96±8.3<br>Baseline Vitamin D level: Placebo (15.62 ng/mL SD 8.69)<br>Vitamin D (15.87 ng/mL SD 8.50)<br>Week 8: Placebo (18.73 ng/mL SD 6.88)<br>Vitamin D (40.02 ng/mL SD 15.33)<br>Change in Vitamin D level: Placebo (3.10 ng/mL SD 4.20)<br>Vitamin D (24.14 ng/mL SD 13.68)<br>Period of follow up: 8 weeks<br>Diagnosis: Adult patients with type 2 diabetes with DPN and vitamin D deficiency | Visual analogue scale<br>Placebo: pre-treatment (5.46 SD 2.13) post-treatment (3.09 SD 2.33) Difference (-2.37 SD 2.2)<br>Vitamin D: pre-treatment (5.74 SD 2.16) post-treatment (2.39 SD 2.09) Difference (-3.34 SD 2.03) | Experimental group received an oral vitamin D 5000 IU (Hi-D 5000) once daily over 8 weeks in addition to the standard treatment (pregabalin 1x75 mg, gabapentin 1x100mg, or amitriptyline 1x25mg; the dose was adjusted according to each patients' symptom) for diabetic neuropathy. | The control group received standard treatment only over the same period. |
| **Davoudi et al** | RCT (five arms), including vitamin D and placebo arms (used in | Total 225 participants. 90 participants selected (45 placebo (5 dropouts) 45 Vitamin D (5 | Neuropathic Pain Severity:<br>Placebo: | Daily 4000 IU oral dosage (four capsules) with 28,000 IU vitamin | Placebo plus usual treatment based on neurologist |
| (2021) | this analysis) | dropouts) | Pre-treatment (47.02 SD 13.7) | D weekly for 12 weeks. | prescribes. |
|  |  | Sex: Placebo (40%F) Vitamin D (55%F) | Post-treatment (45.6 SD 13.4) |  |  |
|  |  | Age: Placebo (56.24+-9.89) | Vitamin D: pre-treatment (49.4 SD 14.7) |  |  |
|  |  | Vitamin D (54.5 +-9.8) | Post-treatment (31.03 SD 14.8) |  |  |
|  |  | Baseline Vitamin D level: Placebo (24.6 ng/mL SD 5.5) |  |  |  |
|  |  | Vitamin D (27.4 ng/mL SD 5.07) |  |  |  |
|  |  | Change in Vitamin D level: Not reported |  |  |  |
|  |  | Period of follow-up: 12 weeks |  |  |  |
|  |  | Diagnosis: Adult patients with type 2 diabetes with neuropathy and vitamin D deficiency. |  |  |  |

footnote: n = number of participants, m = male participants, f = female participants, sd = standard different, iu = internation unit (the amount of a substance that has a certain biological effect)

With respect to study quality, all studies were rated using the GRADE criteria to assess for overall risk-of-bias. One study [26] maintained an over-all low risk of bias by adhering to stringent experimental techniques to minimise bias. Two studies ([27] [28]) were moderate risk-of-bias due to omitting key techniques such as concealment of allocation and effective double blinding. It is not clear what the impact this limitation may have on the observed effect sizes, however sub-par allocation concealment is generally associated with the exaggerated inflation of an effect relative to the true effect [30]. For full summary of quality assessment criteria see table 4 and summary of Cochrane Risk of Bias see figure 2 [below].

**Figure 2:**
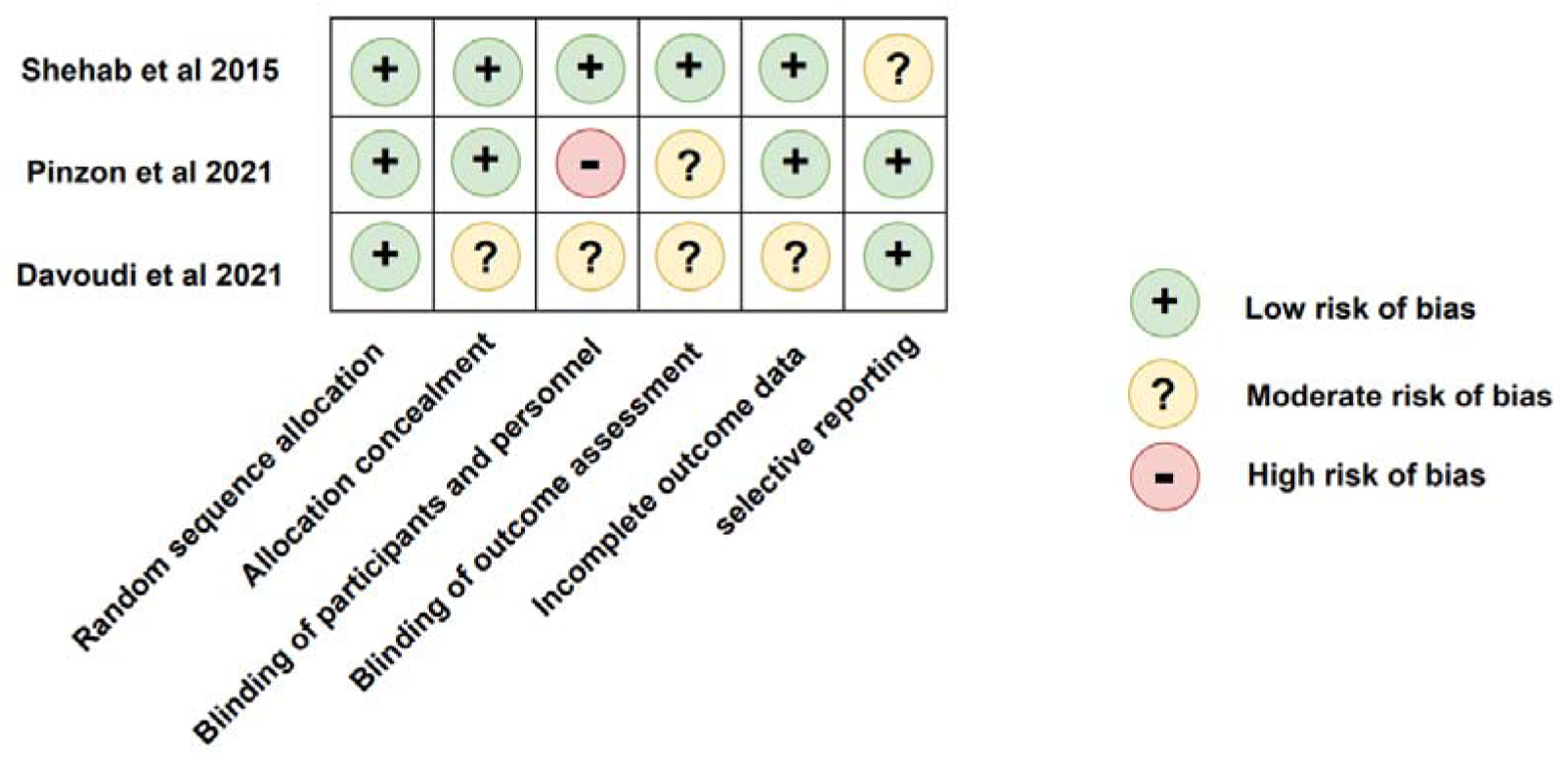
Cochrane risk of bias assessment

**TABLE 4:** Quality Assessment of studies using Cochrane Risk of Bias tool.

| Study | Randomisation<br>-Sequence generation<br>-Allocation concealment (random sequence) | Blinding<br>-Masking of study participants and personnel | Attrition<br>-Outcome data not available |
| --- | --- | --- | --- |
| **Shehab et al (2015)**<br>**Overall: low risk-of-bias** | Did not explicitly state randomised, but participants allocated to groups according to a sequential order generated by the hospital recording system.<br>Concealed opaque envelopes were prepared for all study participants for allocation into the vitamin D treatment group or the placebo group. This was done under the direct<br>control of a single investigator (H.M.).<br>Y – low risk-of-bias | Double-blinded study.<br>The study participants and the outcome assessor were blinded to the treatment allocation until the conclusion of the study.<br>Y low risk-of-bias | Two patients dropped out of the treatment group and 3 out of the placebo group.<br>These dropouts are acknowledged however are not accounted for in the analysis. 95% of participants analysed so considered low risk.<br>No intention to treat<br>N low risk of bias |
| **PINZON ET AL (2021)**<br>**Overall: moderate risk-of-bias** | Randomization was carried out using block randomization with a 1:1 ratio. (allocation of participants may be predictable and result in selection bias when the study groups are unmasked)<br>A randomization list was generated by a statistician not involved with the study, using blocks of 5 stratifications.<br>No concealment of allocation mentioned<br>however allocation performed by an external assessor.<br>Y low risk-of-bias | Complete blinding was considered difficult and not possible. Participants were informed of key elements of the respective intervention and follow-up they were randomized to, but not on information about the treatment and follow-up alternatives in the other group or the study's hypotheses.<br>Complete blinding not performed. Participants were only informed of the intervention for the group they were allocated to.<br>N moderate risk-of-bias | All participants who were lost to follow-up were accounted for in the consort flow diagram.<br>Intention to treat analysis used<br>Y low risk-of-bias |
| **Davoudi et al (2021)**<br>**Overall: moderate risk-of-bias** | Participants were randomly allocated into groups using a random table. No further details given.<br>No concealment of allocation mentioned.<br>Y moderate risk-of-bias | All patients were blinded to receive VD or placebo groups.<br>Single blind<br>No mention of blinding of assessment of outcome<br>Y moderate risk of bias | Toward covering potential dropouts, the sample size increased 25% and reaches to 45 subjects for each group.<br>Less than 95% participants analysed and no ITT analysis performed<br>Patient dropouts are not acknowledged in consort flow diagram and not accounted for in the statistical analysis.<br>N moderate risk-of-bias |

### Meta-analysis

A fixed effects meta-analysis was performed using RevMan 5. All studies were positive [favouring vitamin D] irrespective of choice of outcome measure (Neuropathic Pain Severity (NPS), Neuropathy Symptom Score (NSS), Visual Analogue Scale (VAS)). Standard effect sizes ranged from SMD =-1.03 (95%CI-1.50 to-0.57) [27] to SMD =-0.32 (95%CI-0.79 to 0.16) [28]. The overall pooled effect size was strongly in favour of Vitamin D supplementation (pooled standard effect size =-0.70; 95%CI - 0.95 to-0.45). There was low to moderate between study heterogeneity (Higgins I = 54.9%). See figure 3.

**Figure 3:**
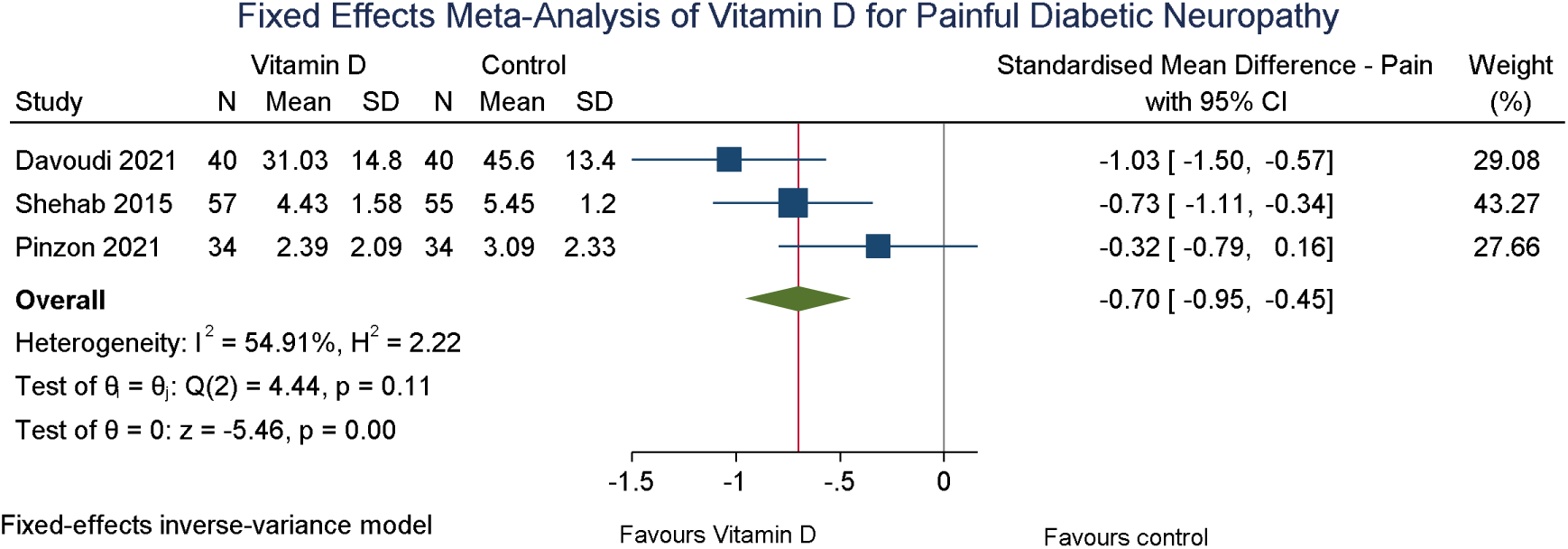
Forest plot of a fixed effect meta-analysis of the effect of Vitamin D on short term outcomes.

The standard effect size of-0.70 was moderate in size, and equivalent to 9.6 (95%CI 13.0 to 6.2) points on the Neuropathic Pain Scale [assuming SD of 13.7, derived from the placebo arm baseline data].

### Test for small study bias

There were three eligible studies, with little variation in sample size, making the Egger forest plot difficult to interpret. The effect of publication and small study bias was therefore difficult to exclude. See figure 4 for Funnel plot. The Egger test was not performed.

**Figure 4:**
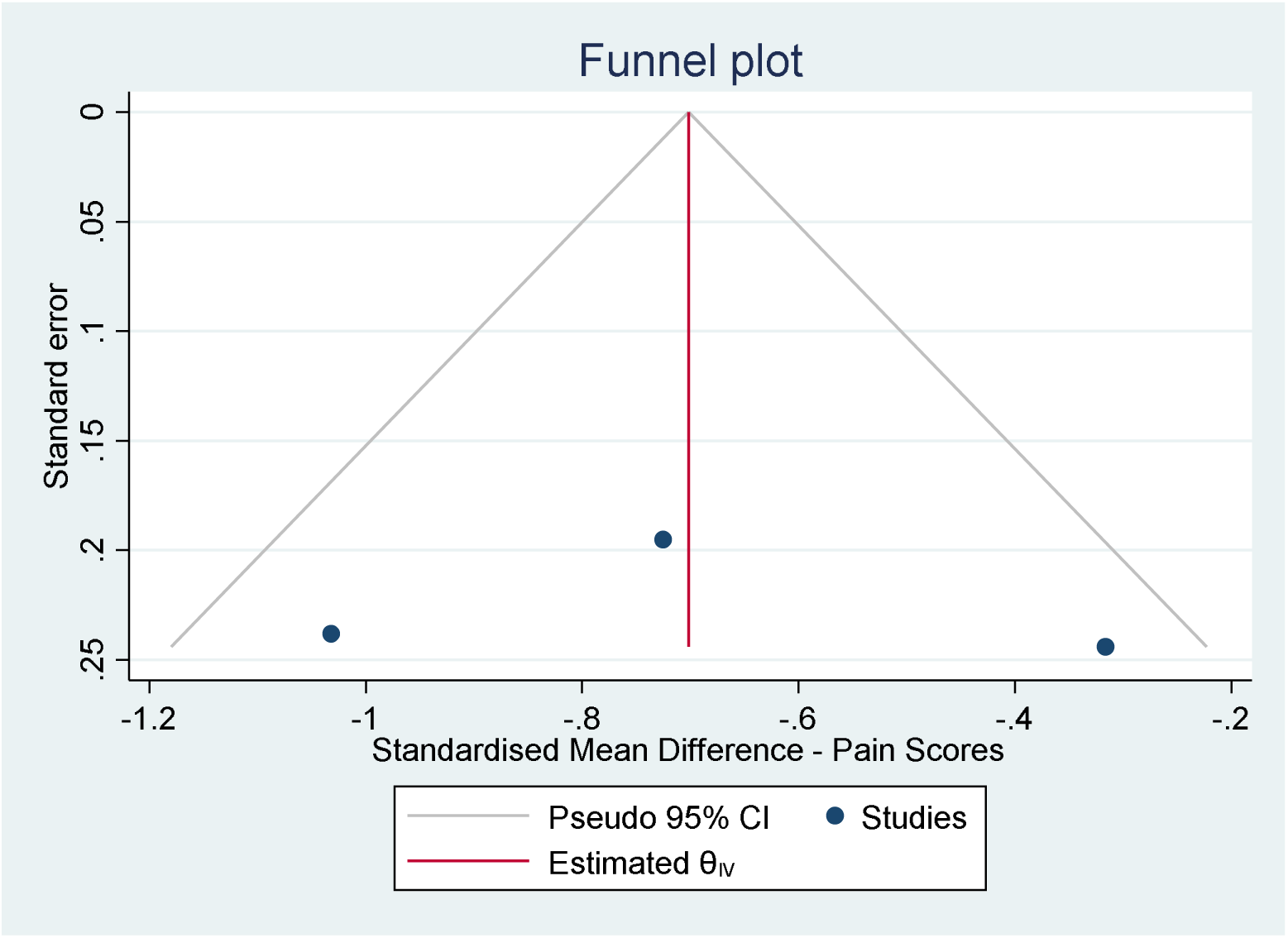
Funnel plot to test for small study bias among included studies

### Selective reporting bias

Two studies were pre-registered and the reported pain outcomes in the final manuscript corresponded with the study protocol [27, 28]. One study was unregistered and there was no available public domain protocol [26].

## DISCUSSION

### Summary of main results

Randomised trials were available to answer the main review question. Three small scale trials showed a reduction in the painful symptoms of peripheral diabetic neuropathy, regardless of the pain outcome used. When the data were pooled, the meta-analysis results demonstrated a significant statistical improvement in reported neuropathic pain by participants. This strongly favours Vitamin D supplementation for the reduction in painful symptoms of diabetic neuropathy compared to control treatment in all trials.

### Agreements and disagreements with other studies or reviews

The review findings are consistent with previous non-experimental studies that have reported Vitamin D as an independent risk factor in peripheral neuropathy in patients with low vitamin D [12]. Non-randomised studies have established that correction of vitamin D deficiency leads to an improvement in symptoms [31], and this is the first time this has been demonstrated in the context of a meta-analysis of RCT data. It is however difficult to establish if the effects of vitamin D supplementation were due to an elevation in the pain threshold, due to improvement of affected nerves, or via improvement in diabetic markers (HbA1c) or a synergy of them all [31–33].

There are plausible biological mechanisms by which this reduction in pain can be explained. Vitamin D is a potent inducer of neurotrophins and neurotransmitters, nerve growth factor (NGF) is such an example [32]. Vitamin D supplementation has been shown to increase nerve growth factor (NGF), a protein required for nerve growth and maintenance in the peripheral nervous system [32].

Epidermal keratinocytes are the primary source of NGF in the skin, lower dermal levels of NGF in patients with diabetic polyneuropathy is associated with neuropathic signs of sensor and autonomic nerve function. This suggests that the diminished levels of NGF in epidermal keratinocytes contribute to the pathogenesis of peripheral neuropathy [32].

Furthermore there is a suggested mechanistic association between neuropathic pain and vitamin D through nociceptive calcitonin gene associated peptide (CGRP)-positive neurons which have been shown to have a distinct vitamin D phenotype alongside hormonally controlled receptor and ligand levels [28, 34].

### Applicability of evidence

Correcting vitamin D levels in patients where there is deficiency is already clinically indicated. However, the observed effects in this preliminary meta-analysis demonstrate the potential added benefit of reduction in painful symptoms of PDN. On top of this, correction of Vitamin D levels is a comparatively low-risk intervention that has also been related to lowering HbA1c levels, reducing insulin resistance and improvement in insulin sensitivity [33, 35]. Therefore, there is the potential for Vitamin D to not only relieve painful symptoms but also improve glycaemic regulation.

Reduction in pain severity and pain disability has many benefits including an improvement in QOL of patients through better outcomes in sleep quality, general activity and mood symptoms [36].

Individuals with reduced pain severity are less likely to use incur healthcare costs through the use of polypharmacy and increased outpatient visits along with increased economic productivity [6, 36].

The widespread use of vitamin D as a adjuvant medication in the control of the symptoms of PDN could therefore be useful in mitigating the high economic and healthcare burden associated with diabetic complications [37].

Of note is that all three of the studies recruited participants who reported a baseline vitamin D deficiency. This remains highly applicable to many patients with painful diabetic neuropathy since Vitamin D deficiency is commonly observed across diabetic populations with one study finding 89% of participants were found to be deficient in Vitamin D with just 9 in 300 participants having sufficient levels of vitamin D [38]. One potential limitation to the findings is the specificity of the results to those patients with an underlying vitamin D deficiency with the outcome effects being as a result of correcting this pre-existing deficiency. This provides limitations to the applicability of these results to non-vitamin D deficient patients with PDN.

It is difficult to make firm conclusions for the applicability of this evidence. However, one important clinical implication is the need for patients with PDN to be routinely tested for vitamin D levels, and to have a low threshold for prescribing Vitamin D for patients at risk of deficiency. Furthermore, the indicated pharmaceutical benefits of vitamin D are significant due to its many perceived advantages over traditional pharmaceutical treatment for diabetic neuropathy. Vitamin D is a relatively low cost, meaning it could be a useful adjunct to existing treatments. With the potential to demonstrate likely cost effectiveness by offsetting the need for prescriptions of expensive analgesic drugs or fewer iatrogenic side effects of high dose analgesic medicines.

According to the NICE treatment pathways for neuropathic pain in adults, vitamin D is not currently recommended due to its limited evidence base [39]. In light of this review, Vitamin D therapy demonstrated a significant effect that might be useful in the formulation of future guidelines.

### Quality of the evidence

The risk of bias assessment indicated a variation in the overall assessment of each study included. There was moderate to high risk of bias for 2 of the studies for the category describing bias in the blinding process. This was typically because of suboptimal blinding process or only single blinding occurring. In the domain bias due to missing outcome data, most studies were found to have a low risk of bias. However, one study was found to have a moderate risk of bias, this was due to failure to perform intention to treat analysis. The impact of the missing data on the observed effects of this study was unclear. In the final domain, bias in the selection of the reported outcome, most papers papers were reported as having a low risk of bias however in one there was an absence of a pre-registered report of trial details thus making it moderate risk of bias. This makes it difficult to make sure that the reported analysis was consistent with the pre-planned analysis.

One limitation of the studies included was the large variation in the prescribed vitamin supplementation ranging from a single weekly oral dose of 50000 IU vitamin D to a daily dose of 5000 IU. Furthermore, analysis was hampered by an overall small sample size (n = 260) over the 3 papers. There is a need to replicate the findings of this review, based on small scale studies, in a large scale randomised controlled trial, with sufficient statistical power. Tests for small study bias using Funnel plots were difficult to interpret, and there is a risk that these results might be overturned by a large-scale trial that is free from publication bias.

### Implications for research

These preliminary results on a relatively small sample size confirm the need for a large fully powered study with a concurrent cost effectiveness analysis to explore whether vitamin D therapy represents good value for money. Mechanisms by which it might demonstrate cost effectiveness include decreased need for polypharmacy, particularly expensive and potentially toxic painkiller medications. This is because the cost of treating diabetic neuropathy is very high and is therefore very important to find the most effective and cost-effective medications [6].

Future research should centre around whether these results are applicable to patients suffering from PDN who do not have underlying vitamin D deficiency and ascertain whether clinical findings are due to the correction of underlying vitamin D deficiency. Furthermore, future research on the proper dosing in addition to the route and duration of treatment is essential to further explore the efficacy and clinical benefits of the use of Vitamin D supplementation for PDN and effectively help advice policy on the dispensation of the drug.

Secondary objectives assessing the effect of vitamin D supplementation on glycaemic control should also be addressed.

## Declaration of interest & contribution

The meta-analysis was carried out by the author Abraham Gilbody. Quality assessment and scrutiny of studies for inclusion was carried out by all authors. Statistical and epidemiological expertise was provided by Dr Joe Gilbody. The authors declare that they have no conflicts of interest, financial support, or grants. We are grateful to Dr Uazman Alam of the University of Liverpool who first interested Dr Abraham Gilbody in this topic as an undergraduate at the University of Liverpool.

## Data Availability

All data produced are available online in this manuscript.

---

**APPENDIX 1:** Criteria for the quality assessment of trials using the Cochrane Risk of Bias tool.

| **Randomization & concealment of allocation:** | **Blinding:** | **Attrition:** |
| --- | --- | --- |
| This will be adequate when patients are allocation into study groups via a method that used numerical randomisation (allocation sequence randomisation). Furthermore, the allocation of study participants must be concealed to both them and trial team members until it is completed. This is to avoid selective enrolment of participants based on the treatment assignment (allocation sequence concealment). | Adequate when double-blinding has occurred, this includes masking of the treatment allocation to both the participants and the trial members. This ensures there is a decreased risk of both deviance from the application of the treatment and the use of non-protocol medication alongside the trial medication (Double blinding). | Adequate when missing outcome data is taken into account in the final analysis and whether the missingness mechanism is linked to the value of the true outcome. Furthermore, the proportion of dropouts must be minimal for the results to be weighted in our analysis. This will avoid bias due to mechanics of the study altering the dropout rate dependant on the intervention given. |

## REFERENCES

1. Walsh, J., et al., Association of diabetic foot ulcer and death in a population-based cohort from the United Kingdom. Diabetic Medicine, 2016. 33(11): p. 1493–1498.

2. Schreiber, A.K., et al., Diabetic neuropathic pain: physiopathology and treatment. World journal of diabetes, 2015. 6(3): p. 432.

3. Sadosky, A., et al., Healthcare utilization and costs in diabetes relative to the clinical spectrum of painful diabetic peripheral neuropathy. Journal of Diabetes and its Complications, 2015. 29(2): p. 212–217.

4. Girach, A., et al., Quality of life in painful peripheral neuropathies: a systematic review. Pain Research and Management, 2019. 2019.

5. Veves, A., M. Backonja, and R.A. Malik, Painful diabetic neuropathy: epidemiology, natural history, early diagnosis, and treatment options. Pain medicine, 2008. 9(6): p. 660–674.

6. Alleman, C.J.M., et al., Humanistic and economic burden of painful diabetic peripheral neuropathy in Europe: A review of the literature. Diabetes Research and Clinical Practice, 2015. 109(2): p. 215–225.

7. Javed, S., et al., Treatment of painful diabetic neuropathy. Therapeutic advances in chronic disease, 2015. 6(1): p. 15–28.

8. Tesfaye, S. and D. Selvarajah, The Eurodiab study: what has this taught us about diabetic peripheral neuropathy? Current diabetes reports, 2009. 9(6): p. 432–434.

9. Control, D. and C.T.R. Group, The effect of intensive treatment of diabetes on the development and progression of long-term complications in insulin-dependent diabetes mellitus. N Engl j Med, 1993. 329(14): p. 977–986.

10. Rosenberger, D.C., et al., Challenges of neuropathic pain: focus on diabetic neuropathy. Journal of Neural Transmission, 2020. 127(4): p. 589–624.

11. Ziegler, D. and V. Fonseca, From guideline to patient: a review of recent recommendations for pharmacotherapy of painful diabetic neuropathy. Journal of Diabetes and its Complications, 2015. 29(1): p. 146–156.

12. Shehab, D., et al., Does Vitamin D deficiency play a role in peripheral neuropathy in Type 2 diabetes? Diabetic medicine, 2012. 29(1): p. 43–49.

13. Shillo, P., et al., Reduced vitamin D levels in painful diabetic peripheral neuropathy. Diabetic Medicine, 2019. 36(1): p. 44–51.

14. Basit, A., et al., Vitamin D for the treatment of painful diabetic neuropathy. BMJ Open Diabetes Research and Care, 2016. 4(1).

15. Higgins, J.P., et al., Cochrane handbook for systematic reviews of interventions. 2019: John Wiley & Sons.

16. Liberati, A., et al., The PRISMA statement for reporting systematic reviews and meta-analyses of studies that evaluate health care interventions: explanation and elaboration. Journal of clinical epidemiology, 2009. 62(10): p. e1–e34.

17. Royle, P. and R. Milne, Literature searching for randomized controlled trials used in Cochrane reviews: rapid versus exhaustive searches. International journal of technology assessment in health care, 2003. 19(4): p. 591–603.

18. Lefebvre, C., E. Manheimer, and J. Glanville, Chapter 6: Searching for studies In Higgins JPT & Green S.(Eds.), Cochrane handbook for systematic reviews of interventions, version 5.1. 0 (updated March 2011). The Cochrane Collaboration. 2011.

19. Hariton, E. and J.J. Locascio, Randomised controlled trials—the gold standard for effectiveness research. BJOG: an international journal of obstetrics and gynaecology, 2018. 125(13): p. 1716.

20. Melzack, R. and J. Katz, The McGill Pain Questionnaire: appraisal and current status. 2001.

21. Perez, C., et al., Validity and reliability of the Spanish version of the DN4 (Douleur Neuropathique 4 questions) questionnaire for differential diagnosis of pain syndromes associated to a neuropathic or somatic component. Health and quality of life outcomes, 2007. 5(1): p. 1–10.

22. Egger, M., et al., Bias in meta-analysis detected by a simple, graphical test. Bmj, 1997. 315(7109): p. 629-634.

23. Raghav, K.P.S., et al., From protocols to publications: a study in selective reporting of outcomes in randomized trials in oncology. Journal of Clinical Oncology, 2015. 33(31): p. 3583.

24. Riley, R.D., J.P. Higgins, and J.J. Deeks, Interpretation of random effects meta-analyses. Bmj, 2011. 342.

25. Higgins, J.P. and S.G. Thompson, Quantifying heterogeneity in a meta-analysis. Statistics in medicine, 2002. 21(11): p. 1539–1558.

26. Shehab, D., et al., Prospective evaluation of the effect of short-term oral vitamin D supplementation on peripheral neuropathy in type 2 diabetes mellitus. Medical Principles and Practice, 2015. 24(3): p. 250–256.

27. Davoudi, M., et al., The synergistic effect of vitamin D supplement and mindfulness training on pain severity, pain-related disability and neuropathy-specific quality of life dimensions in painful diabetic neuropathy: a randomized clinical trial with placebo-controlled. Journal of Diabetes & Metabolic Disorders, 2021. 20(1): p. 49–58.

28. Pinzon, R.T., V.O. Wijaya, and V. Veronica, The Benefits of Add-on Therapy of Vitamin D 5000 IU to the Vitamin D Levels and Symptoms in Diabetic Neuropathy Patients: A Randomized Clinical Trial. Journal of Pain Research, 2021. 14: p. 3865.

29. Mehta, S., et al., Effectiveness of Empagliflozin With Vitamin D Supplementation in Peripheral Neuropathy in Type 2 Diabetic Patients. Cureus, 2021. 13(12).

30. Schulz, K.F., et al., Empirical evidence of bias: dimensions of methodological quality associated with estimates of treatment effects in controlled trials. Jama, 1995. 273(5): p. 408–412.

31. Bell, D.S., Reversal of the symptoms of diabetic neuropathy through correction of vitamin D deficiency in a type 1 diabetic patient. Case Reports in endocrinology, 2012. 2012.

32. Carlson, A.N. and A.M. Kenny, Is vitamin D insufficiency associated with peripheral neuropathy? The Endocrinologist, 2007. 17(6): p. 319–325.

33. Dalgård, C., et al., Vitamin D status in relation to glucose metabolism and type 2 diabetes in septuagenarians. Diabetes care, 2011. 34(6): p. 1284–1288.

34. Tague, S.E. and P.G. Smith, Vitamin D receptor and enzyme expression in dorsal root ganglia of adult female rats: modulation by ovarian hormones. Journal of chemical neuroanatomy, 2011. 41(1): p. 1–12.

35. Pittas, A.G., et al., The role of vitamin D and calcium in type 2 diabetes. A systematic review and meta-analysis. The Journal of Clinical Endocrinology & Metabolism, 2007. 92(6): p. 2017–2029.

36. Sadosky, A., et al., Burden of illness associated with painful diabetic peripheral neuropathy among adults seeking treatment in the US: results from a retrospective chart review and cross-sectional survey. Diabetes, metabolic syndrome and obesity: targets and therapy, 2013. 6: p. 79.

37. Stedman, M., et al., Cost of hospital treatment of type 1 diabetes (T1DM) and type 2 diabetes (T2DM) compared to the non-diabetes population: a detailed economic evaluation. BMJ open, 2020. 10(5): p. e033231.

38. Lee, J.-I., et al., Serum 25-hydroxyvitamin D concentration and arterial stiffness among type 2 diabetes. Diabetes research and clinical practice, 2012. 95(1): p. 42–47.

39. Nice, C.f.C.P.a., Neuropathic pain: the pharmacological management of neuropathic pain in adults in non-specialist settings [Internet]. 2013.

